# SARS-CoV-2 Inactivation Potential of Metal Organic Framework Induced Photocatalysis

**DOI:** 10.1101/2020.10.01.20204214

**Authors:** Jason Ornstein, Ray O.K. Ozdemir, Anne Boehme, Farid Nouar, Christian Serre, Daniel N. Ackerman, Vicki L. Herrera, Joshua L. Santarpia

## Abstract

As the world recovers from the lockdown imposed by the Severe Acute Respiratory Syndrome Coronavirus 2 (SARS-CoV-2) pandemic, returning to shared indoor spaces is considered a formidable risk. It is now clear that transmission of SARS-CoV-2 is driven by respiratory microdroplets expelled by infected persons, which can become suspended in the air. Several layering technologies are being explored to mitigate indoor transmission in the hopes of re-opening business, schools and transportation systems. Here we coupled the water adsorptive and photocatalytic capacity of novel Metal Organic Frameworks (MOFs) to demonstrate the capture and inactivation of SARS-CoV-2. Discussion is given on the methods of analysis and the differences between the photocatalytic activity of several MOFs, and the difference between MOF induced photocatalysis and ultra violet photolysis of SARS-CoV-2. Our results are intended to provide support to industry looking for alternative methods secure indoor spaces.

## Editorial

The ability to prevent the accumulation, through ongoing passive inactivation, of Severe Acute Respiratory Syndrome Coronavirus 2 (SARS-CoV-2) on surfaces that may pose ongoing infection risk may be critical in indoor confided spaces, especially when occupied by infected patients. Previous studies have shown that dehumidification systems can collect and spread SARS-COV-2.[1] Photocatalysis on surfaces where respiratory droplets are likely to accumulate is one possible method for rendering dehumidification systems safe. We propose using the desiccant Metal Organic Frameworks (MOFs), which can also function as a photocatalyst to break down adsorbed guest molecules, may lead to viral inactivation, for hospital dehumidification systems.

The COVID-19 pandemic has demanded a large number of airborne infection isolation rooms (AIIR), which has put enormous stress on our medical infrastructure. Because closed-loop air flow dehumidification systems can turn any room into a negative pressure isolation room, researchers from the Republic of South Korea were able to demonstrate that this system could be used as an AIIR during the MERS outbreak.[2] In parallel, the microporous crystalline Ti dicarboxylate MOF MIL-125(Ti)-NH_2_, [3] has been shown to improve dehumidification efficiency with heat pumps, [4][5] and Ti-MOFs are known to be efficient photocatalysts.[6] Given these properties, we tested the ability of this MOF to inactivate SARS-COV-2 through UV-irradiation.

MIL-125(Ti)-NH_2_ is built up from Ti_8_O_8_(OH)_4_ oxoclusters connected via amino-terephtalate ligands to produce a microporous cubic 3D porous architecture. It can be produced as nano- or micro-particles under ambient pressure and solvothermal conditions and exhibits a good stability in the presence of water while being suitable for a wide range of potential applications. The MOF was produced, activated and characterized according to previous studies. The fine yellowish powder was then mixed with deionised (DI) water at 5 weight percent. 1 mL of the solution was dispersed evenly on a one inch by one inch reflective stainless steel coupon. The solution was then allowed to dry until all of the water evaporated. SARS-CoV-2 virus (BEI_ USA-WA1/2020) cultivated in Vero-E6 cells and enumerated by median tissue culture infectious dose (TCID50) assay to be ∼1×10^5^ TCID50/mL. The MOF nano particles were then exposed to 100 µL of SARS-CoV-2 culture and exposed to UV-C light for 30 minutes. A control consisting of a stainless steel coupon and 100 µL of SARS-CoV-2 culture was also maintained for 30 minutes. As a point of comparison, UV-C light is currently used to sanitize face masks where the practice is to expose the mask for much longer durations to ensure viral inactivation.[7]

After the exposure, the sample was analyzed for remaining viral titer by TCID50 and RNA content by Reverse Transcriptase PCR (RT-PCR) targeting the E gene of SARS-CoV-2 [8]. RT-PCR indicate only the presence of viral RNA, not the activity of the virus. Although significant reduction in viral RNA and inactivation of virus, by TCID50, was observed it could not be separated from the impacts of UV-C alone due to the choice of a reflective coupon substrate (Figure 1). It is likely that the reflectivity of the coupon increased the exposure intensity for the control coupon, compared to the MOF coated ones. However, compared to other photoactive MOF materials such as the iron(III) carboxylate MIL-127(Fe), also referred to as PCN-250(Fe), which was tested in the same experiment,[9] MIL-125(Ti)-NH_2_ appears to exhibit additional inactivation (Figure 1).

**Figure 1.**
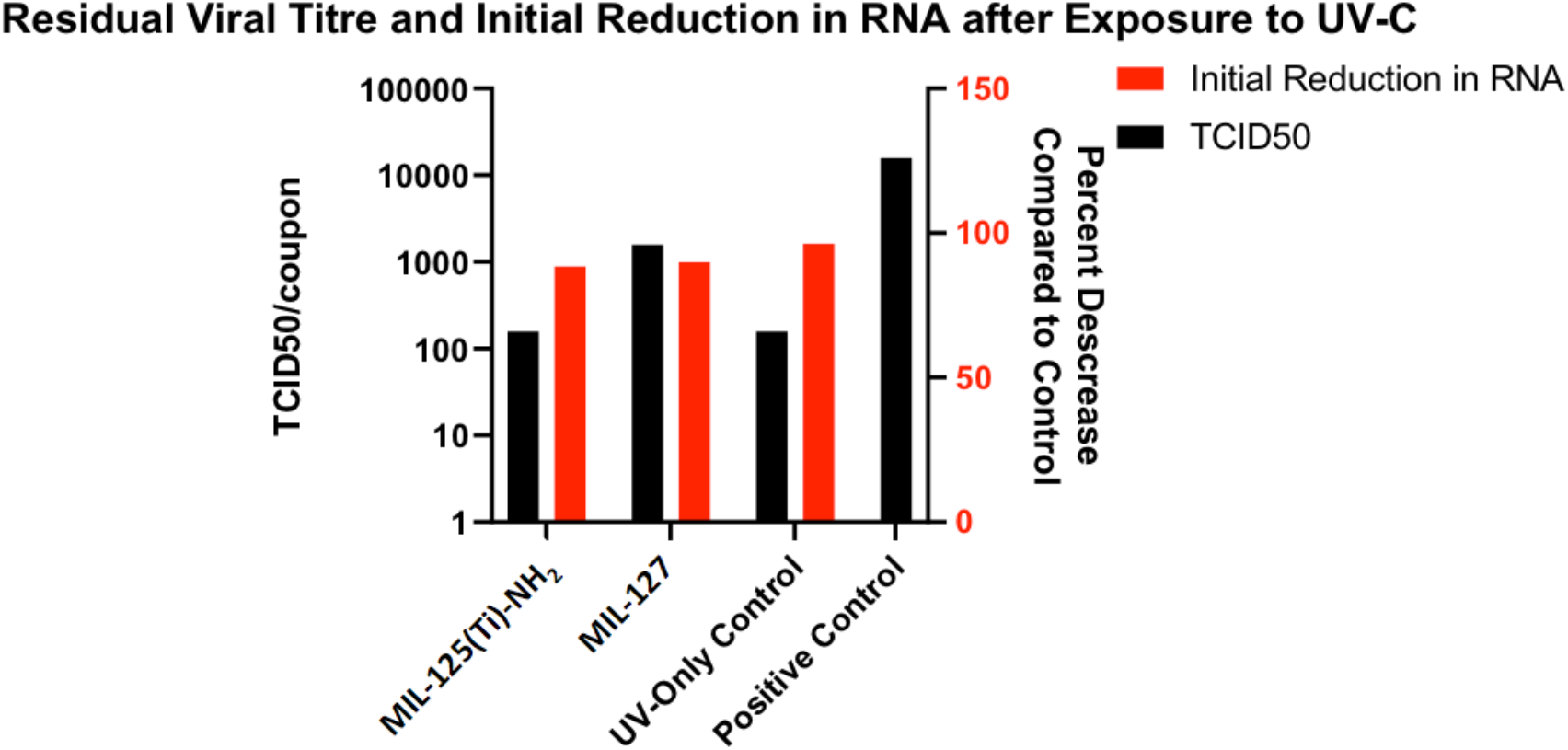
PCR and TCID50 test results showing SARS-CoV-2 Inactivation Coupon treated with MIL-125(Ti)-NH_2_ demonstrates greater inactivation (by TCID50) when exposed to UV-C radiation than MIL-127(Fe), although reduction in RNA is identical, indicating RNA damage is not the primary means of inactivation.

It has been proposed previously that photocatalytic virus inactivation can be achieved through three distinct pathways, which prevent SARS-CoV-2 from initiating an infection: (i) nucleic acid DNA damage,[10] (ii) alteration to spike proteins or recognition proteins on the surface,[11] and (iii) virus’ lipid membrane damage.[12] Direct UV damage to nucleic acid occurs at the wavelengths absorbed by the RNA in the germicidal UV region and can also be referred to as photolysis.[13] Photocatalysis, or oxidative degradation of organic material, on the other hand, can target the recognition proteins and the lipid membrane, while leaving some genetic material intact. The role of photocatalytic processes versus photolysis could not be determined by this study; however, this preliminary data indicates that photocatalytic activity was observed for MIL-125(Ti)-NH_2_, since greater inactivation by an order of magnitude was observed for MIL-125(Ti)-NH_2_ than for MIL-127(Fe), even though initial reduction in RNA was similar.

MIL-177-HT is a microporous MOF built up from ca. 1.1 nm Ti oxide chains connected via 3,3′,5,5′-tetracarboxydiphenylmethane ligands to produce a nanoporous honeycomb porous architecture. [14] The presence of the infinite Ti oxide chains, associated with a very high oxo/Ti ratio of 1.5, creates a continuous photoconductor pathway enabling longer charge separation under photo excitation. This provides significant advantages in terms of reactive species generation, which benefits to photocatalytic remediation applications. It can also be produced as ≈150 nm (length) nanorods under ambient pressure solvothermal conditions; it also exhibits a very good stability in the presence of water while being promising for a wide range of applications. [15][16]

The MOF was produced, activated and characterized according to previous studies.[17] The fine white powder was then mixed with DI water at 5 weight percent. 1 mL of the solution was dispersed evenly on a one inch by one inch [non-reflective coupon, as well as MIL-125(Ti)-NH_2_ on a separate coupon and a control coupon with no MOF material. Coupons were inoculated with virus, as defined above. The sample was exposed to florescent light for 30 minutes. Most of the spectral power for florescent lights occur in the visible range.[18]

After the exposure, the sample was analyzed for remaining viral titer by TCID50. Noteworthy, MIL-177-HT achieved 50% virus inactivation (Figure 2). Room light alone had no effect on SARS-CoV-2 during our testing, and this is believed to be the first demonstration of the use of room light, which unlike UV-C, is not harmful to humans, to drive MOF photocatalysis of SARS-CoV-2, or any virus. The difference between MIL-125(Ti)-NH_2_ and MIL-177-HT could arise from the photoconductive character of the latter.[17] This MOF exhibits a much longer life time of the separated charge under irradiation in comparison with MIL-125(Ti)-NH_2_. MIL-177-HT is thus the first of its kind to exhibit photoconductivity stemming from the titanium oxide building block.

**Figure 2.**
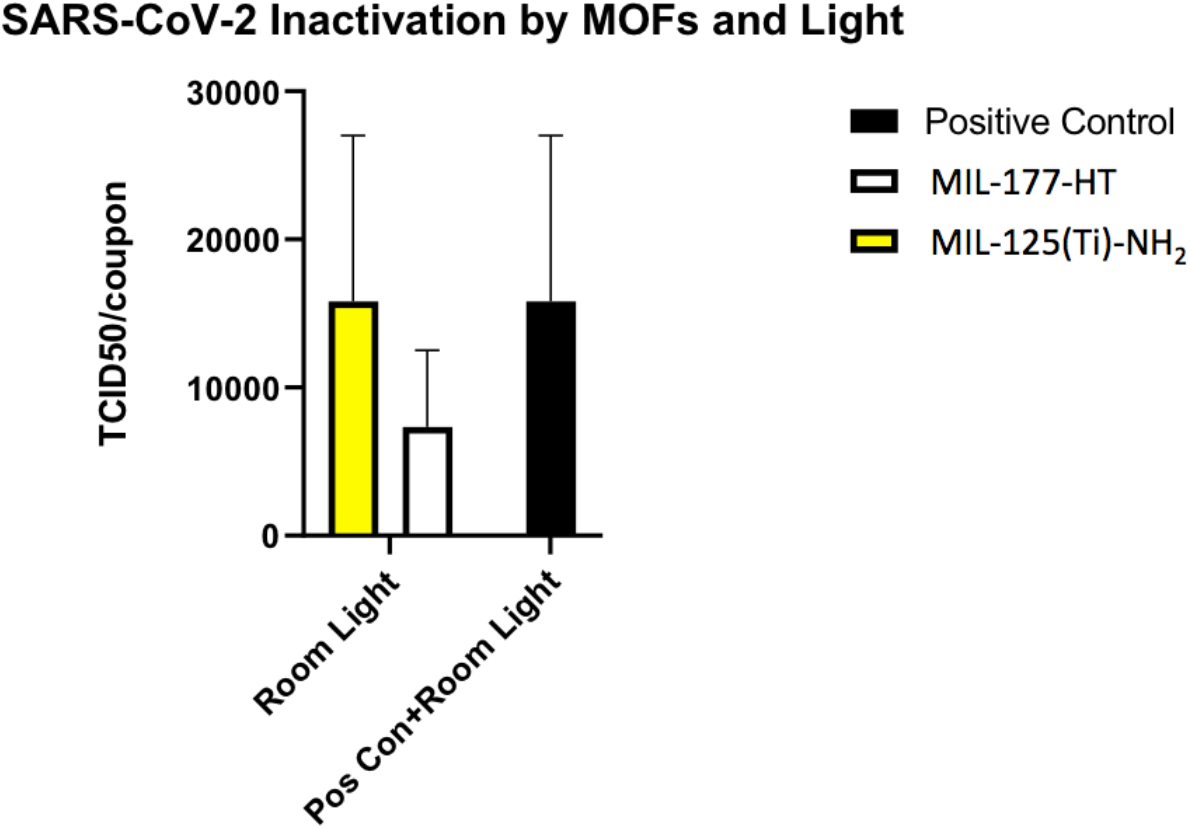
TCID50 test results showing SARS-COV-2 Inactivation in Room Light- MIL-177-HT MOF-coated surfaces shows 50% reduction in viral titer after a 30 minute exposure to room light, compared to the control and MIL-125(Ti)-NH_2_ coated coupons.

The results of these preliminary studies therefore suggest more testing should be done on the efficiency of MOF photocatalysis of SARS-CoV-2, and other viruses and biological contaminants. Based on these preliminary findings, the coupling of incredible water adsorptive capacity with a photocatalytic capability of MOFs could be a practical risk reduction solution in dehumidification systems in hospitals. This technology could also be applied in other confined spaces and to other negative air pressure systems such as air purifiers and breathing masks.

## Data Availability

All data generated or analysed during this study are included in this paper. Material characterization information including the crystallinity raw data is available via contributing authors.

## Acknowledgements

This approach benefits from work being done by framergy, Inc., under funding from the US Environmental Protection Agency, where Metal Organic Frameworks are being used to photocatalytically degrade per- and polyfluoroalkyl substances in an aqueous medium (Contract Number 68HERC20C0055).

## Materials and methods

### Synthesis of MIL-125(Ti)-NH_2_

MIL-125(Ti)-NH_2_ was synthesized using 300 g of 2-aminoterephthalic acid (ligand), 150 mL Titanium (IV) isopropoxide 98+% (Ti(iPrO)_4_), 8.1 L DMF, 900 mL Methanol. A 10 L reactor was heated to 120°C. 300 g of ligand was dissolved in 4.5 L of DMF while stirring and heating in a glass container until a uniform solution was obtained. This solution was added to the heated reactor. Then, in a 500 mL glass flask, 150 mL of Titanium (IV) isopropoxide was dissolved in another 3.5 mL of DMF. To the reactor, 900 mL of methanol was added, along with 100 mL of DMF. This caused the ligand to completely dissolve. The dissolved titanium isopropoxide/DMF solution was added to the mixture in the reactor and the temperature of the oil surrounding the reactor, was increased to 150 °C. The mixture was stirred and heated in the reaction at 150°C for 48 hours where after it was cooled to room temperature. The yellow solid was then filtered using a Nutsche, washed in methanol and filtered once more, leaving a dry yellow powder.

**Figure 3.**
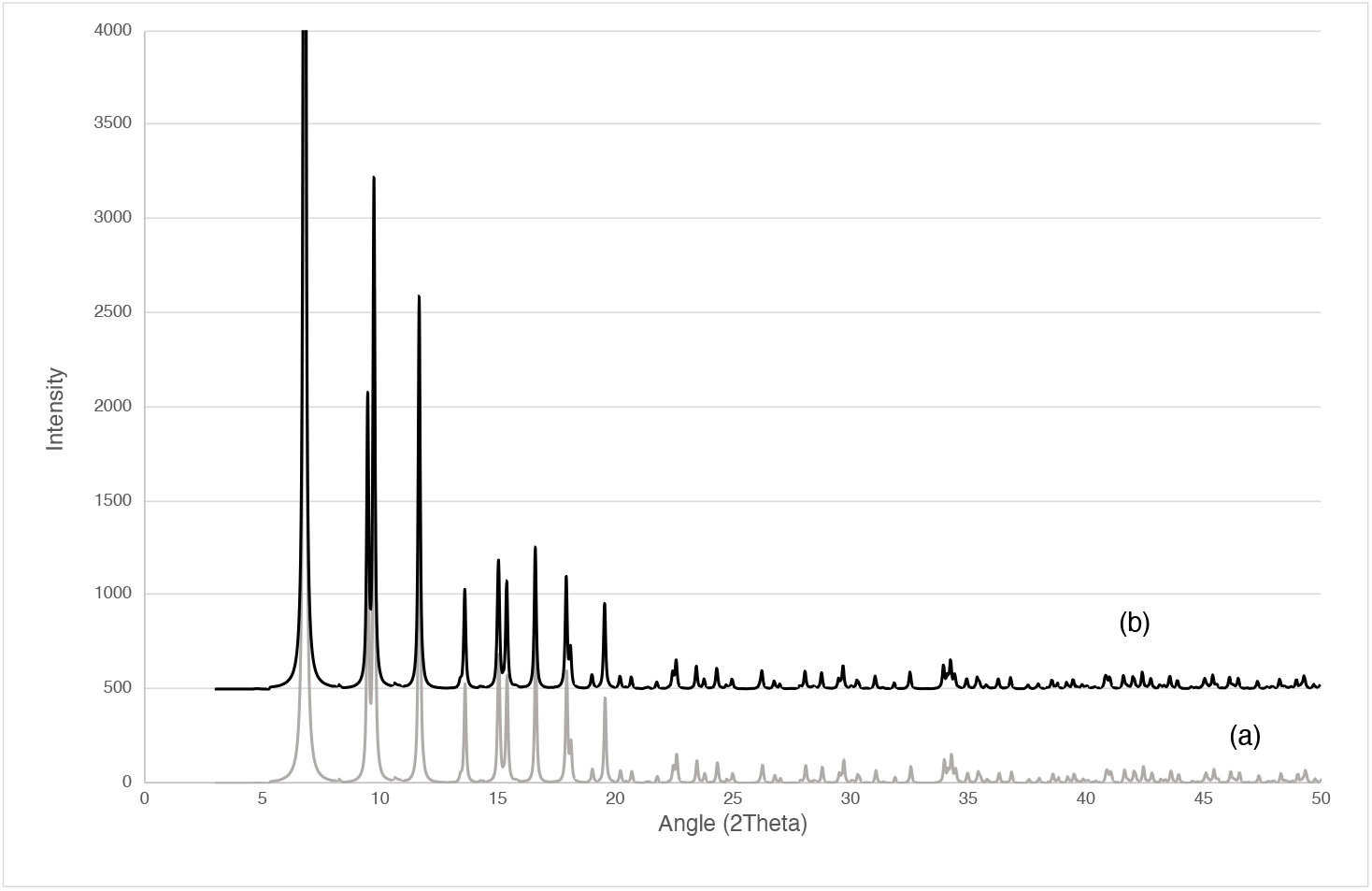
PXRD patterns of a) calculated MIL-125(Ti)-NH_2_ and b) experimental MIL-125(Ti)-NH_2_

### Synthesis of MIL-177-HT

MIL-177-HT was synthesized using a 25 mL round bottom flask where 200 mg of H4mdip with 10 mL of formic acid was added and stirred at room temperature until it was a uniform solution. Following that, 400 µL of Titanium (IV) isopropoxide 98+% (Ti(iPrO)4) was added drop wise using a micropipette. The reaction was heated under reflux for 24 hours and then cooled to room temperature. Then, the white solid was filtered using vacuum filtration and washed with ethanol, where after it was filtered once more. MIL-177-HT was prepared by grounding 200 mg of the MOF into a fine powder, which was transferred to a flat glass dish, and dispersed uniformly. It was then heated in an oven at 280 °C for 12 hours, forming a white powder.

**Figure 4.**
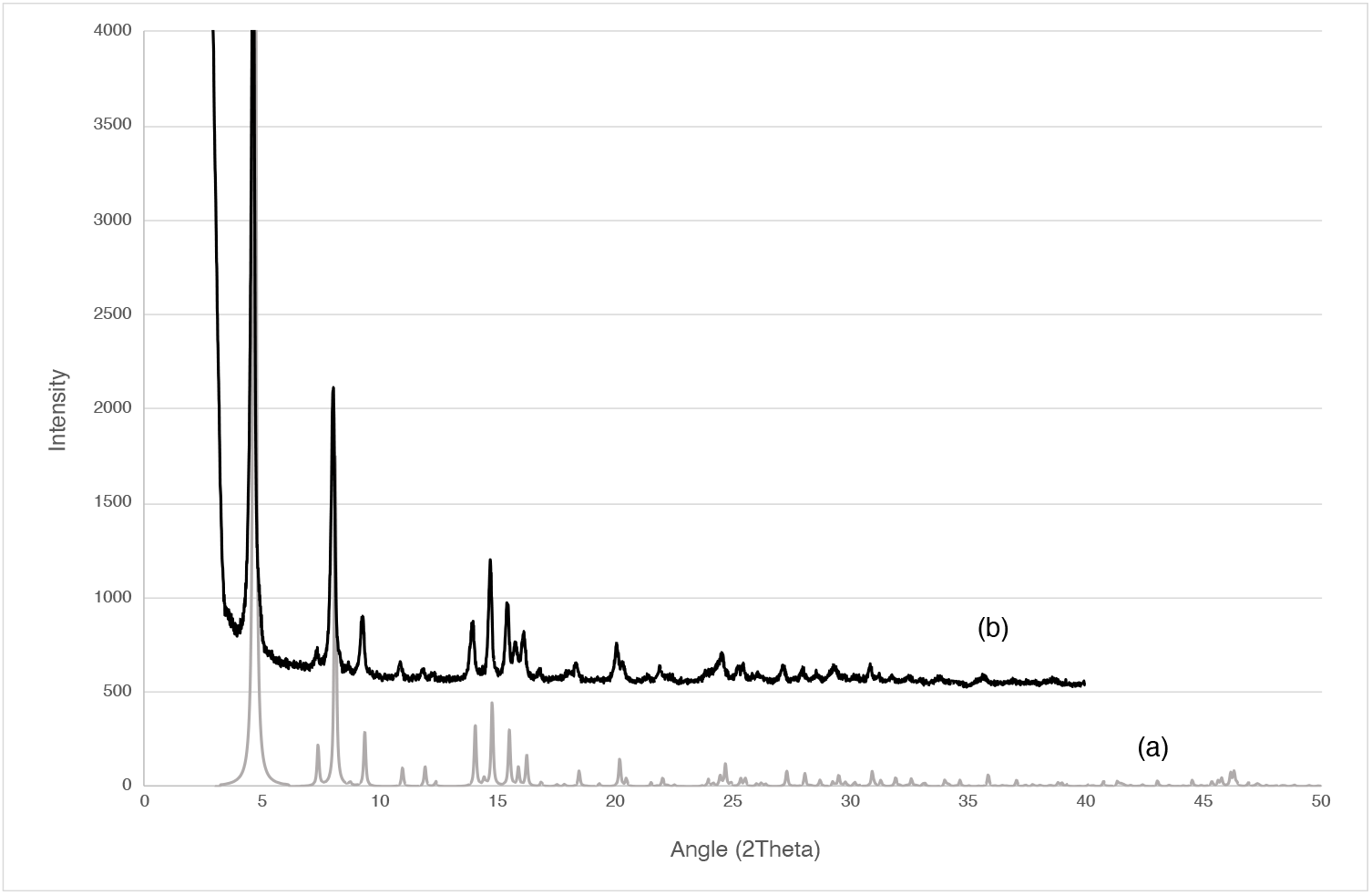
PXRD patterns of a) calculated MIL-177-HT and b) experimental MIL-177-HT

### Synthesis of MIL-127(Fe)

MIL-127(Fe) was synthesized using 6 L of DMF, 3 L acetic acid, 90 g of H4ABTC (azobenzene-tetracarboxylic acid), and 270 g of iron nitrate. A 10 L reactor was heated to 150°C. First, the 6 L DMF was combined with the 90 g H4ABTC. The solution was stirred at room temperature until it was a uniform solution. The solution was added to the 10 L reactor. Then, in another beaker, the acetic acid (3 L) and the iron nitrate (270 g) was combined and stirred until a uniform solution was obtained. The mixture was then added to the 10 L reactor as well. The final solution was heat at 150°C for 12 hours, where after it was cooled to room temperature. The dark brown solid crystals were then filtered using a Nutsche, washed in methanol and filtered once more, leaving a dry dark red powder.

**Figure 5.**
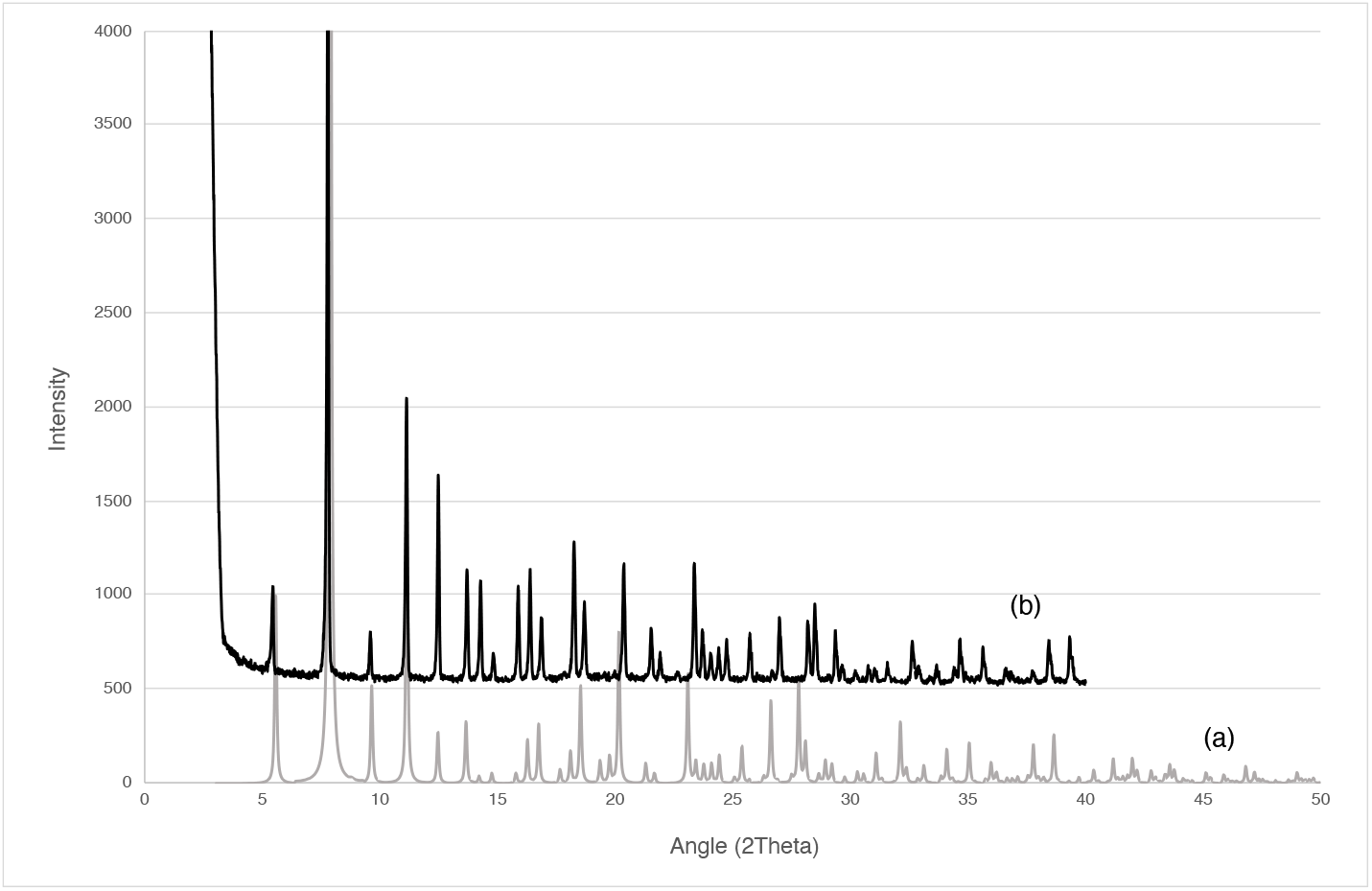
PXRD patterns of a) calculated MIL-127(Fe) and b) experimental MIL-127(Fe)

